# Algorithmic implementation of pancreatic cancer staging guidelines: comparison with a retrieval-augmented large language model

**DOI:** 10.64898/2026.06.30.26356912

**Authors:** Atsushi Komaba, Akitomo Amakawa, Ryota Tozuka, Junichi Sato, Koudai Fujihara, Mizuki Emoto, Shin Sawada, Shigeki Kasai, Kasane Sakamoto, Karen Shimura, Yuki Johno, Kazunori Nakamoto, Shintaro Ichikawa, Hisashi Johno

**Author notes:** **Corresponding Author** Hisashi Johno, MD, PhD, Department of Diagnostic Radiology, Faculty of Medicine, University of Yamanashi, 1110 Shimokato, Chuo, Yamanashi 409-3898, Japan. **Author contributions Atsushi Komaba:** Methodology, Software, Validation, Formal analysis, Investigation, Resources, Data curation, Writing – review & editing, Visualization. **Akitomo Amakawa:** Methodology, Validation, Formal analysis, Data curation, Writing – review & editing, Visualization. **Ryota Tozuka:** Investigation, Writing – review & editing, Project administration. **Junichi Sato:** Investigation, Data curation, Writing – review & editing. **Koudai Fujihara:** Investigation. **Mizuki Emoto:** Investigation. **Shin Sawada:** Investigation. **Shigeki Kasai:** Investigation. **Kasane Sakamoto:** Investigation. **Karen Shimura:** Investigation. **Yuki Johno:** Validation, Writing – review & editing. **Kazunori Nakamoto:** Methodology, Resources, Writing – review & editing, Supervision, Funding acquisition. **Shintaro Ichikawa:** Supervision, Writing – review & editing. **Hisashi Johno:** Conceptualization, Methodology, Validation, Formal analysis, Writing – original draft, Writing – review & editing, Visualization, Supervision, Project administration.

## Abstract

**Purpose:** To implement a comprehensive knowledge-based algorithm (KBA) for pancreatic cancer staging based on the current Japanese guidelines and to evaluate its performance as a clinical decision support system in comparison with a retrieval-augmented large language model (LLM) system.

**Materials and methods:** A KBA covering TNM classification, stage classification, and resectability classification was implemented as a web application. The correctness of the system outputs was exhaustively verified for all possible inputs. Subsequently, six non-board-certified radiologists performed pancreatic cancer staging for 12 simulated cases with imaging findings under three conditions: unassisted, LLM-assisted, and KBA-assisted. Staging accuracy and staging time were compared among the three conditions using pairwise proportion *z*-tests and Welch’s *t*-tests, respectively.

**Results:** In the comparative experiment, staging accuracy was 81.9%, 80.6%, and 98.6% in the unassisted, LLM-assisted, and KBA-assisted conditions, respectively. Mean staging time was 229.2, 401.9, and 196.2 s, respectively. The KBA-assisted condition showed higher accuracy than both the unassisted and LLM-assisted conditions (both *p*<0.001). Staging time was longer in the LLM-assisted condition than in the other two conditions (both *p*<0.001).

**Conclusion:** A comprehensive KBA for pancreatic cancer staging based on the current Japanese guidelines was implemented and exhaustively verified. In a preliminary comparative experiment, KBA assistance improved staging accuracy without increasing staging time, whereas LLM assistance increased staging time without improving staging accuracy. These findings suggest that verified KBA systems may be feasible and useful for clinical tasks governed by explicit guideline-based rules.

**Secondary Abstract:** A comprehensive knowledge-based algorithm (KBA) for pancreatic cancer staging was implemented and exhaustively verified. In a preliminary comparative experiment involving six radiologists and 12 simulated cases, KBA assistance improved staging accuracy without increasing staging time, whereas retrieval-augmented large language model (LLM) assistance increased staging time without improving staging accuracy.

## 1. Introduction

Clinical decision support systems (CDSS) are broadly categorized into two types: knowledge-based systems, which algorithmically implement clinical guidelines, and non-knowledge-based systems, which generate outputs using machine learning [1,2]. The key characteristics of these two approaches are summarized in Table 1. Traditionally, knowledge-based systems have been considered advantageous in that their underlying logic can be explicitly verified and corrected by humans, and have therefore been recommended for clinical use [1–3].

**Table 1.** Comparison of key characteristics between non-knowledge-based and knowledge-based clinical decision support systems (CDSS).

|  | **Non-knowledge-based CDSS** | **Knowledge-based CDSS** |
| --- | --- | --- |
| **Mechanism** | Learning-based (e.g., LLMs) | Rule-based (e.g., Yes/No decision rules) |
| **Guideline usage** | Used as training data or prompts | Algorithmically implemented |
| **User input** | Unstructured (e.g., free text) | Structured (e.g., checklist) |
| **Reasoning process** | Opaque (black-box) | Explicit and auditable |
| **Output correctness** | Cannot be guaranteed | Can be guaranteed |
| **Clinician verification** | Required for every output | Primarily required for inputs |

Before such knowledge-based approaches became widely established in clinical practice, large language model (LLM)-based systems, particularly those incorporating retrieval-augmented generation (RAG), have rapidly gained attention as a new form of CDSS [4,5]. These systems are considered non-knowledge-based, as they generate outputs probabilistically based on training data rather than through predefined rules [4–7].

Cancer staging is a key application of CDSS. Recent studies have increasingly reported the potential utility of LLMs, particularly for pancreatic cancer staging based on Japanese guidelines [7–11]. However, these studies have consistently highlighted inherent limitations of non-knowledge-based systems, including unavoidable errors (hallucinations) and the requirement for clinician verification of every output. In contrast, a comprehensive knowledge-based algorithm (KBA) implementing the current Japanese pancreatic cancer staging guidelines [12], including the complex resectability classification, has not yet been established.

The aims of this study were threefold: (1) to implement a KBA for pancreatic cancer staging based on the current Japanese guidelines; (2) to exhaustively verify that the KBA system produces correct outputs for all possible inputs; and (3) to conduct a preliminary comparative experiment in which radiologists performed pancreatic cancer staging under three conditions—without assistance (unassisted), with assistance from a retrieval-augmented LLM system (LLM-assisted), and with assistance from the proposed KBA system (KBA-assisted)—and to compare staging accuracy and staging time.

## 2. Materials and methods

Fig. 1 illustrates the conceptual differences between LLM-assisted and KBA-assisted pancreatic cancer staging.

**Fig. 1.**
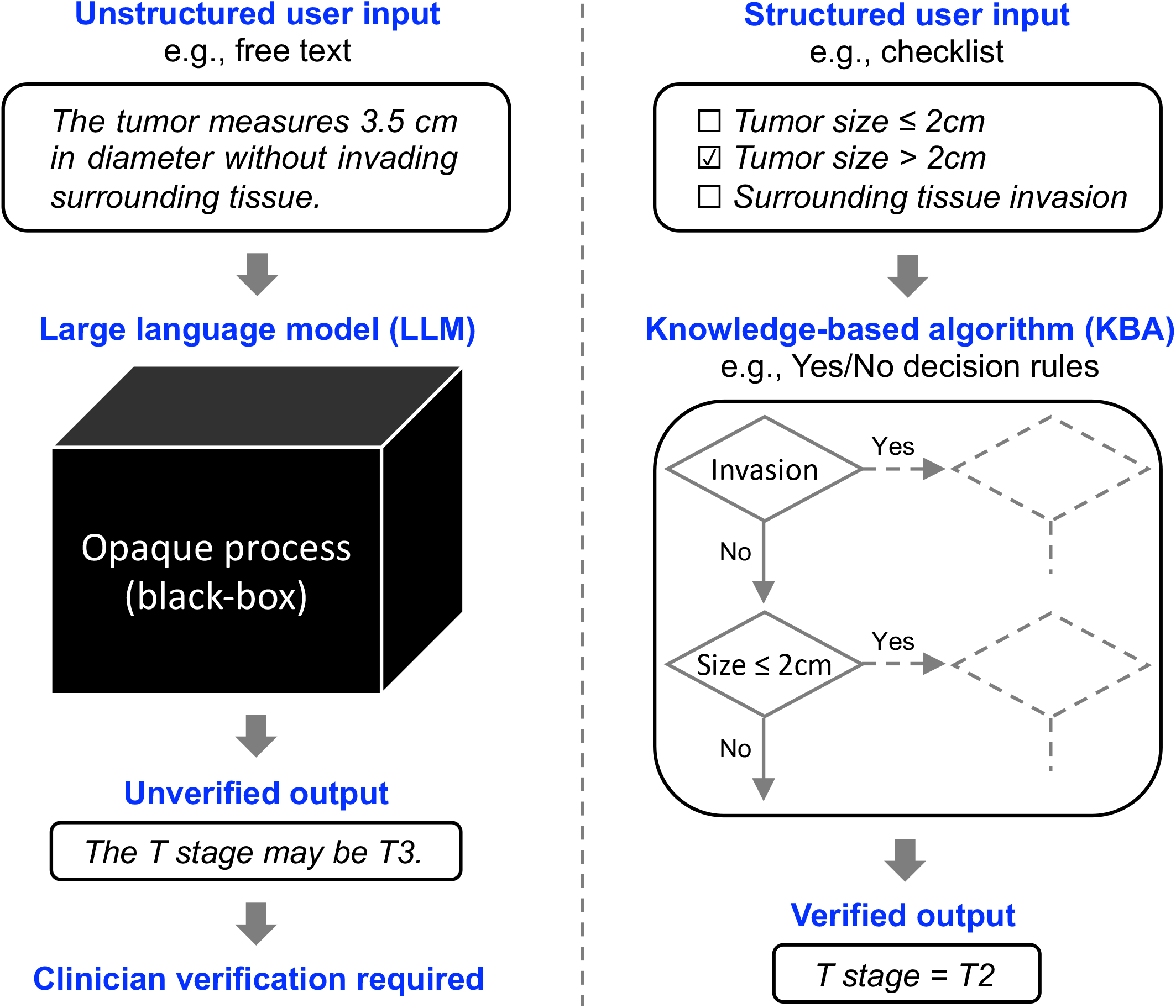
Conceptual comparison of LLM-assisted and KBA-assisted pancreatic cancer staging. In the LLM-assisted condition, imaging findings are provided to the LLM system as unstructured user input (e.g., free text). The system generates outputs through an opaque (black-box) process, and their correctness cannot be guaranteed; therefore, clinician verification is required for every output. In the KBA-assisted condition, imaging findings are provided to the KBA system as structured user input (e.g., a checklist). The system generates outputs through explicit and auditable rules (e.g., Yes/No decision rules). If all possible outputs have been verified in advance, every output produced during use is guaranteed to be correct, provided that the input is correct. LLM = large language model, KBA = knowledge-based algorithm.

### 2.1 KBA system for pancreatic cancer staging

The logical structure of the latest pancreatic cancer staging guidelines in Japan, namely the eighth edition of the Japanese Classification of Pancreatic Carcinoma [12], was analyzed by one board-certified radiologist and two engineers, one of whom is also a physician and the other a medical student. Based on this analysis, a KBA system was developed to determine T classification, N classification, M classification, overall stage, and resectability classification by implementing guideline-defined decision logic. The system operates on structured inputs and produces deterministic outputs. Clinical variables derived from imaging findings, including tumor location, lymph node metastases, distant metastases, and local invasion factors, are predefined and selected through an English-language graphical user interface (Fig. 2). Once the required inputs are provided, the algorithm is automatically executed, and the corresponding staging results are displayed. The system was implemented as a JavaScript-based web application using React and Vite, and is publicly available at https://tnm-en.vercel.app/.

**Fig. 2.**
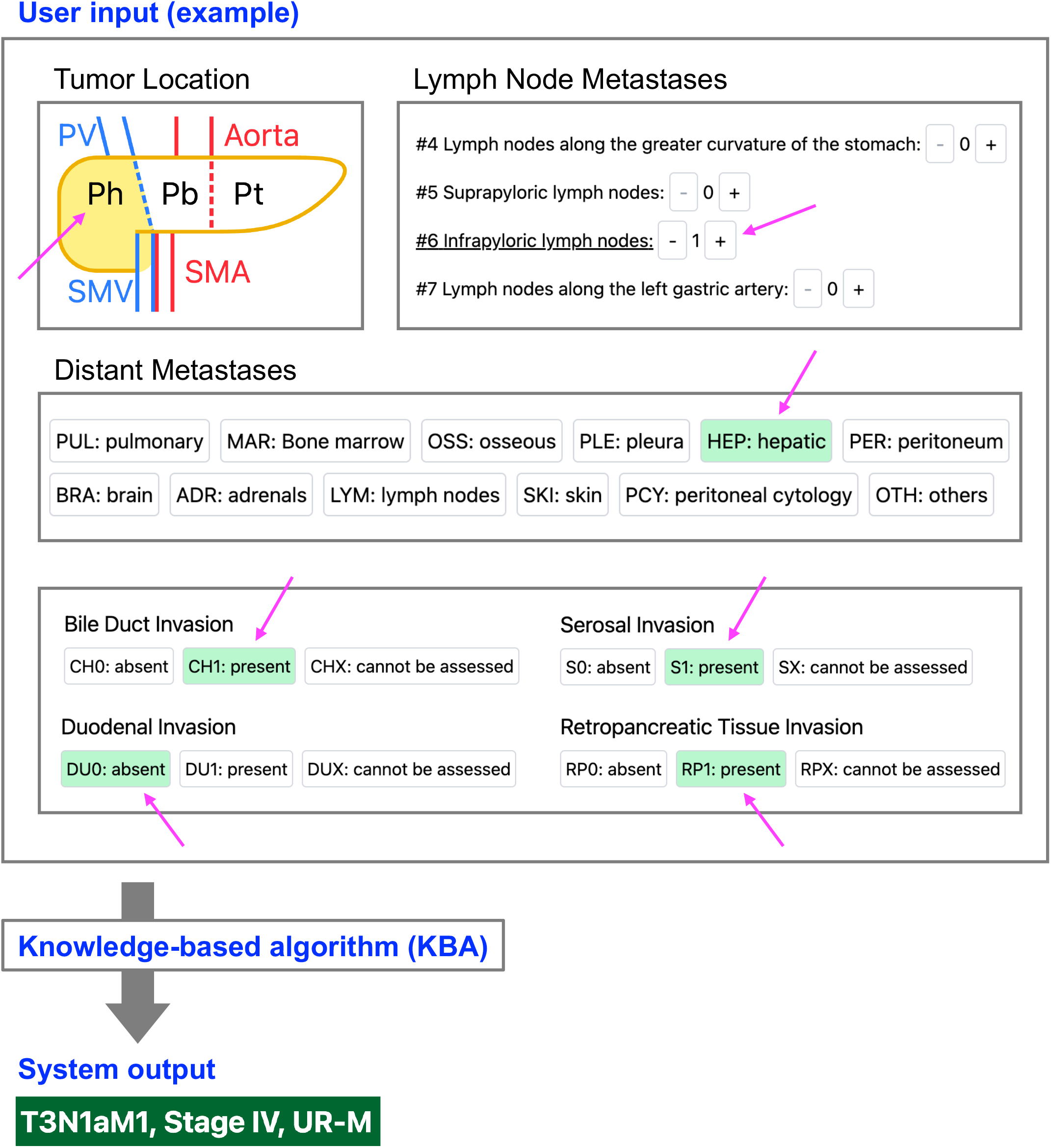
Graphical user interface of the KBA system for pancreatic cancer staging. Users access the web application (https://tnm-en.vercel.app/) and provide structured inputs by selecting predefined variables, including tumor location, lymph node metastases, distant metastases, and local invasion factors. Upon completion of the required fields, a deterministic JavaScript-based algorithm is automatically executed, and the corresponding staging information is immediately displayed as the system output. KBA = knowledge-based algorithm.

Since the system produces deterministic and reproducible outputs for the finite set of possible inputs, all input-output combinations were exhaustively listed, and the correctness of each output was verified by two board-certified radiologists, one board-certified gastroenterologist, and the two engineers (Supplementary Material 1, Section 1, including Supplementary Fig. 1 and Supplementary Tables 1–4). The correctness of all outputs was confirmed by consensus among the evaluators.

### 2.2 Retrieval-augmented LLM-based staging system

Portions of the outputs of a retrieval-augmented LLM system for pancreatic cancer staging, previously reported in [11], were used in the present experiment. Among the multiple LLMs evaluated in that study, outputs generated by Gemma-3 27B (Google) were selected. The system employed BAAI/bge-base-en-v1.5 (Beijing Academy of Artificial Intelligence) as the embedding model for RAG.

### 2.3 Comparative experiment

Six non-board-certified radiologists with 1–5 years of radiology experience performed pancreatic cancer staging for 12 simulated cases with imaging findings described in English under three conditions: without assistance (unassisted), with assistance from the retrieval-augmented LLM system (LLM-assisted), and with assistance from the KBA system (KBA-assisted); staging accuracy and staging time were then evaluated for each condition. The 12 cases were taken from [11]. To maintain a relatively uniform level of difficulty, only cases with T3 or T4 classification were selected and renumbered from 1 to 12 (Supplementary Material 1, Section 2.1). In the LLM-assisted condition, the participants were provided with pre-generated LLM outputs (Supplementary Material 1, Section 2.2) rather than direct access to the LLM system. Although the same 12 cases were repeatedly evaluated under the three conditions, the experiments were conducted at 1-month intervals to reduce the influence of radiologists’ short-term memory. In addition, the assignment of the three conditions across the first, second, and third experimental sessions was balanced such that each radiologist-condition-session combination contained four cases (Supplementary Material 1, Section 2.3 and Supplementary Table 5). The raw experimental results used for statistical analysis are provided in Supplementary Material 2.

### 2.4 Statistical Analysis

Statistical analyses were performed in Python using the packages `statsmodels` (version 0.14.5), `SciPy` (version 1.13.1), `pandas` (version 2.2.2), `NumPy` (version 1.26.4), and `matplotlib` (version 3.9.2). Staging was considered correct only when all staging components (T classification, N classification, M classification, overall stage, and resectability classification) were correctly classified.

To evaluate factors associated with staging correctness, logistic regression analysis was performed. For each observation *i* ∈ {1,2, …,216}, corresponding to a unique condition-radiologist-case combination, let *Y*_*i*_ ∼ Bernoulli(*p*_*i*_) be the Bernoulli random variable representing staging correctness, where *p*_*i*_ = *P*(*Y*_*i*_ = 1) is the probability of correct staging. We considered the logistic regression model

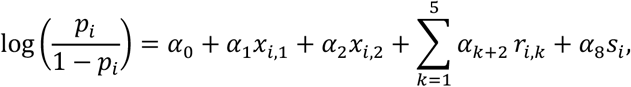

where *α*_0_, *α*_1_, …, *α*_8_ ∈ ℝ are regression coefficients, *x*_*i*,1_, *x*_*i*,2_ ∈ {0,1} are dummy explanatory variables for the conditions, defined as

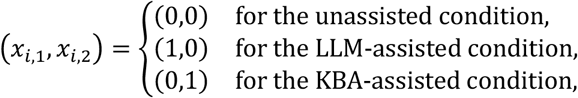

*r*_*i*,1_, *r*_*i*,2_, …, *r*_*i*,5_ ∈ {0,1} are the dummy explanatory variables for the radiologists, defined as

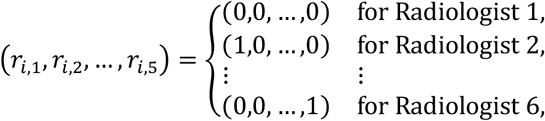

and *s*_*i*_ ∈ {1,2,3} is the explanatory variable representing the experimental session number. The coefficients were estimated by maximizing the likelihood function

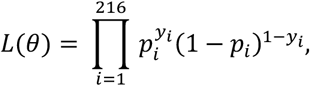

where *θ* = (*α*_0_, *α*_1_, …, *α*_8_) and *y*_*i*_ ∈ {0,1} is the observed realization of *Y*_*i*_. Let 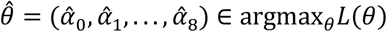 and 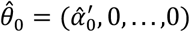, where 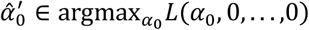. The likelihood ratio statistic was given by 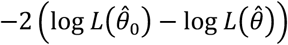 and its *p*-value under the null hypothesis *α*_1_ = ⋯ = *α*_8_ = 0 was calculated. Here, 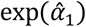 and 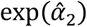 represent the odds ratios for the LLM-assisted and KBA-assisted conditions relative to the unassisted condition, respectively. Similarly, 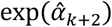 represents the odds ratio for Radiologist *k* + 1 relative to Radiologist 1 for *k* ∈ {1, …,5}, and 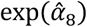 represents the multiplicative change in the odds of correct staging per experimental session. The 95% confidence intervals (CI) and *p*-values for the odds ratios were also calculated.

Condition-specific staging accuracies and their 95% CI were calculated using the Wilson score interval. Pairwise comparisons of staging accuracy between conditions were performed using two-sample proportion *z*-tests, and *p*-values for the null hypothesis of equal proportions between conditions were calculated.

To evaluate factors associated with the time required for staging, analysis of variance (ANOVA) based on the linear regression model

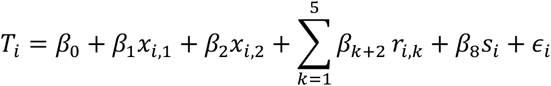

was performed, where *T*_*i*_ denotes the staging time for observation *i* ∈ {1,2, …,216}, *β*_0_, *β*_1_, …, *β*_8_ ∈ ℝ are regression coefficients, and *ϵ*_*i*_ is the error term. The explanatory variables were defined as described above. The unassisted condition and Radiologist 1 were treated as the reference categories for the condition and radiologist variables, respectively. Type II ANOVA was performed for the fitted linear regression model. For each explanatory factor (condition, radiologist, and session), sum of squares, *F*-statistics, and corresponding *p*-values were calculated to test the null hypothesis that all regression coefficients associated with that factor were equal to zero.

Pairwise comparisons of staging time between conditions were performed using Welch’s *t*-tests. Mean differences, 95% CI, and *p*-values for the null hypothesis of equal mean staging times between conditions were calculated.

## 3. Results

The logistic regression model showed association between the explanatory variables and correct pancreatic cancer staging (likelihood ratio test, *p*<0.001) (Table 2). Compared with the unassisted condition, the KBA-assisted condition was associated with higher odds of correct staging (OR, 21.44; 95% CI, 2.53 to 181.52; *p*=0.005), whereas the LLM-assisted condition showed little difference (OR, 0.89; 95% CI, 0.35 to 2.26; *p*=0.813). Experimental session number was also associated with correct staging (OR, 3.14 per session increase; 95% CI, 1.65 to 5.97; *p*<0.001). Among the radiologists, Radiologist 3 showed lower odds of correct staging than Radiologist 1 (OR, 0.20; 95% CI, 0.05 to 0.83; *p*=0.027), whereas the remaining radiologists showed smaller differences.

**Table 2.** Logistic regression analysis of factors associated with correct pancreatic cancer staging. Unassisted condition and Radiologist 1 were used as the reference categories.

|  | Odds ratio (95% CI) | *p*-value |
| --- | --- | --- |
| **LLM-assisted vs. Unassisted** | 0.89 (0.35, 2.26) | 0.813 |
| **KBA-assisted vs. Unassisted** | 21.44 (2.53, 181.52) | 0.005 |
| **Radiologist 2 vs. Radiologist 1** | 1.00 (0.21, 4.87) | 1.000 |
| **Radiologist 3 vs. Radiologist 1** | 0.20 (0.05, 0.83) | 0.027 |
| **Radiologist 4 vs. Radiologist 1** | 2.32 (0.36, 14.87) | 0.375 |
| **Radiologist 5 vs. Radiologist 1** | 1.00 (0.21, 4.87) | 1.000 |
| **Radiologist 6 vs. Radiologist 1** | 1.44 (0.27, 7.71) | 0.672 |
| **Session (per session increase)** | 3.14 (1.65, 5.97) | <0.001 |

LLM = large language model, KBA = knowledge-based algorithm, CI = confidence interval.

Staging accuracy was 81.9% (95% CI, 71.5% to 89.1%) in the unassisted condition, 80.6% (95% CI, 70.0% to 88.0%) in the LLM-assisted condition, and 98.6% (95% CI, 92.5% to 99.8%) in the KBA-assisted condition (Fig. 3, left panel). Pairwise proportion tests showed little difference between the unassisted and LLM-assisted conditions (*p*=0.831), whereas the KBA-assisted condition showed higher accuracy than both the unassisted (*p*<0.001) and LLM-assisted (*p*<0.001) conditions.

**Fig. 3.**
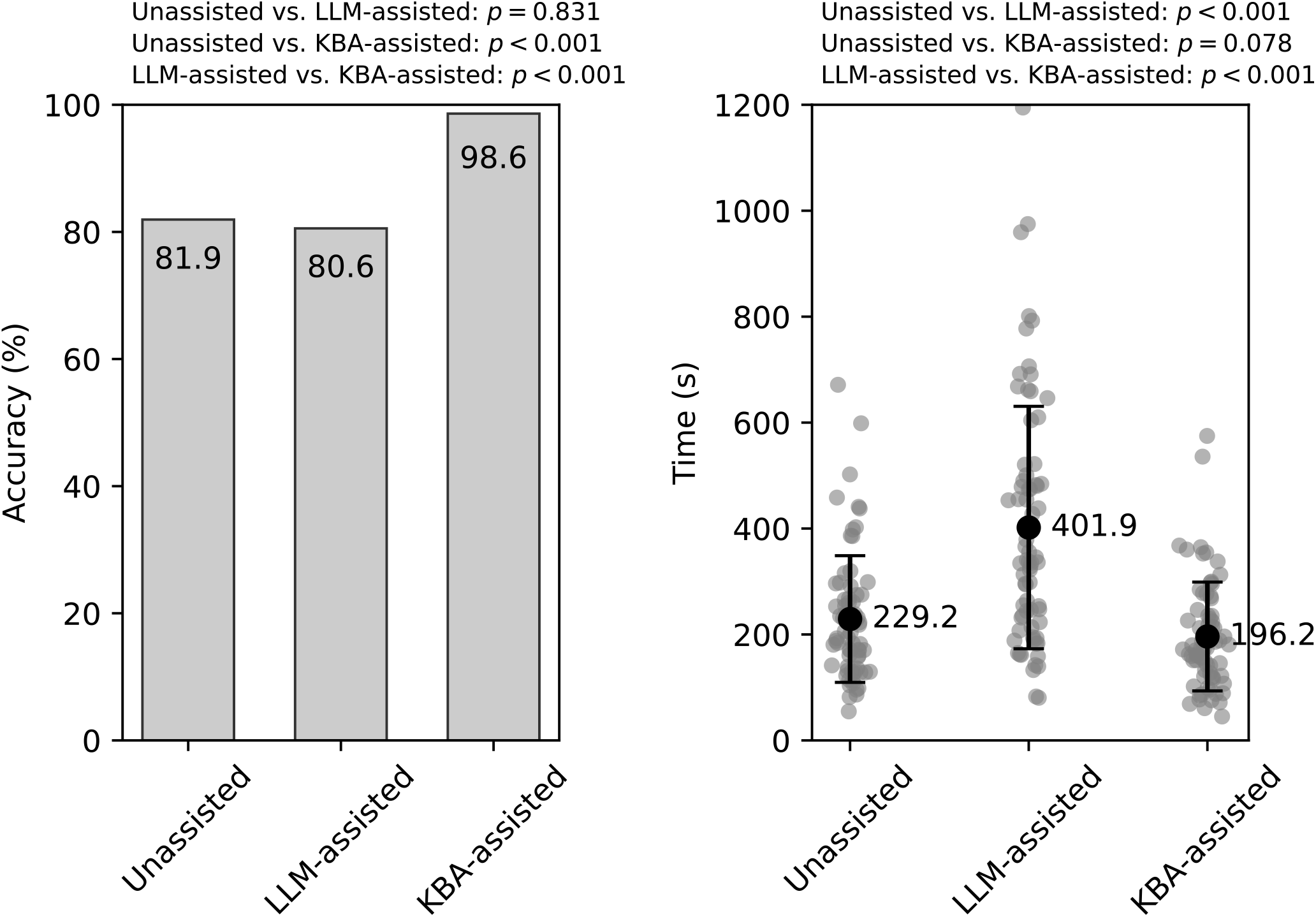
Staging accuracy and staging time across experimental conditions. Left: staging accuracy for each condition. Bars indicate the proportion of correct classifications. Right: staging time for each condition. Individual observations are shown as gray scatter points. Mean values are shown as black circles with error bars representing ±1 standard deviation. *P*-values from pairwise proportion tests (left) and pairwise Welch’s *t*-tests (right) are shown. LLM = large language model, KBA = knowledge-based algorithm.

Type II ANOVA showed associations between staging time and condition (*F*=46.17, *p*<0.001), radiologist (*F*=9.41, *p*<0.001), and experimental session number (*F*=34.06, *p*<0.001) (Table 3). Staging time (mean ± standard deviation) was 229.2 ± 119.6 s, 401.9 ± 228.9 s, and 196.2 ± 102.7 s in the unassisted, LLM-assisted, and KBA-assisted conditions, respectively (Fig. 3, right panel). Pairwise Welch’s *t*-tests showed little difference between the unassisted and KBA-assisted conditions (mean difference, 33.0 s; 95% CI, −3.8 to 69.7 s; *p*=0.078). In contrast, staging time in the LLM-assisted condition exceeded that in both the unassisted condition (mean difference, 172.7 s; 95% CI, 112.4 to 233.0 s; *p*<0.001) and the KBA-assisted condition (mean difference, 205.7 s; 95% CI, 147.0 to 264.4 s; *p*<0.001).

**Table 3.** Type II ANOVA for factors associated with staging time.

| **Explanatory factor** | **Sum of squares** | **df** | ***F*-statistic** | ***p*-value** |
| --- | --- | --- | --- | --- |
| **Condition** | 1,757,415 | 2 | 46.17 | <0.001 |
| **Radiologist** | 895,403 | 5 | 9.41 | <0.001 |
| **Session** | 648,254 | 1 | 34.06 | <0.001 |
| **Residual** | 3,940,016 | 207 | - | - |

## 4. Discussion

In this study, we implemented a KBA for pancreatic cancer staging based on the current Japanese guidelines and exhaustively verified its correctness for all possible inputs. We also conducted a preliminary comparative experiment in which radiologists performed pancreatic cancer staging with and without assistance from the proposed KBA system and a retrieval-augmented LLM system. In this experiment, KBA assistance improved staging accuracy without increasing staging time, whereas LLM assistance increased staging time without improving staging accuracy.

The principal contribution of this study was the establishment of a comprehensive KBA system for pancreatic cancer staging and its exhaustive verification. To our knowledge, such a system covering the entire staging process, including the resectability classification, has not previously been established. Several recent studies have instead explored the potential utility of LLMs for pancreatic cancer staging [7–11]. However, these studies have consistently highlighted the unavoidable risk of hallucinations, an inherent limitation of LLM systems that does not arise in a correctly implemented KBA system.

In the preliminary comparative experiment, LLM assistance increased staging time without improving staging accuracy. This may reflect the need for radiologists to individually verify LLM-generated outputs, which may be incorrect. In contrast, KBA assistance improved staging accuracy without increasing staging time, possibly because radiologists could rely on the outputs of the exhaustively verified system. Nevertheless, staging accuracy in the KBA-assisted condition did not reach 100%. The single incorrect case was attributable to incorrect data entry by the radiologist. Thus, even a correctly implemented and verified KBA system may produce an incorrect result when erroneous information is provided as input.

This study has several limitations. First, although the proposed KBA system was based on the Japanese pancreatic cancer staging guidelines, it was implemented in English, and the comparative experiment was conducted entirely in English, including the imaging findings. Therefore, staging accuracy and staging time may differ from those observed in a Japanese-language clinical setting. Second, the experiment was preliminary and involved a limited number of radiologists and simulated pancreatic cancer cases rather than cases derived from actual clinical practice. Consequently, caution is warranted when extrapolating these findings to routine clinical practice.

Future work should include development and evaluation of a Japanese-language version of the proposed system using actual pancreatic cancer imaging findings described in Japanese. It will also be important to investigate the applicability of KBA systems to other guideline-based clinical tasks and to identify situations in which KBA systems may be preferable to LLM-based systems.

## Data Availability

Data used and analyzed during this study are provided in Supplementary Materials 1 and 2.

## Acknowledgements

This study was partially supported by JSPS KAKENHI Grant Number JP24K06686.

## 5. Declaration

### Ethics approval

This study was approved by the Institutional Review Board of the Faculty of Medicine, University of Yamanashi (Approval No. 2967; expedited review, June 24, 2025).

### Informed consent

The requirement for informed consent was waived by the Institutional Review Board because the study involved voluntary participation of physicians and was considered to pose minimal risk.

### Data availability

Data used and analyzed during this study are provided in Supplementary Materials 1 and 2.

