## Supplementary Material 1 for "Algorithmic implementation of pancreatic cancer staging guidelines: comparison with a retrieval-augmented large language model"


#### 1 Verification of the Algorithmic Staging System

The purpose of this section is to exhaustively verify the correctness of the outputs generated by our pancreatic cancer staging system (`https://tnm-en.vercel.app/`) for all possible inputs.

##### 1.1 Exhaustive Enumeration of Input-Output Combinations

In the graphical user interface, the user selects the tumor location (head, body, or tail; Supplementary Fig. 1). If none is selected, it is recorded as unselected. Regional lymph nodes are determined based on the selected location; if multiple locations are selected, the first is treated as the primary site. The number of lymph node metastases at stations #1–#18 is then entered; if not assessable, it is recorded as NX. The presence of metastases is specified by selecting the corresponding anatomical sites from predefined categories: PUL (pulmonary), MAR (bone marrow), OSS (osseous), PLE (pleura), HEP (hepatic), PER (peritoneum), BRA (brain), ADR (adrenals), LYM (lymph nodes), SKI (skin), PCY (peritoneal cytology), and OTH (others).

Supplementary Table 1 lists all possible combinations of input variables (tumor location, lymph node metastases, and distant metastases) along with their corresponding system outputs (N and M classifications). Supplementary Table 2 similarly lists all possible combinations of input variables (primary tumor category, local invasion factors, and largest tumor diameter) and the corresponding system outputs (T classification).

Based on the T, N, and M classifications, the stage classification is uniquely determined. Supplementary Table 3 shows all possible TNM classification combinations and their corresponding system outputs (stage classifications).

Finally, system outputs (resectability classifications) for all possible combinations of input variables (vascular invasion and distant metastasis) are exhaustively summarized in Supplementary Table 4.

##### 1.2 Expert Validation of System Outputs

All input-output combinations for T, N, and M classifications, as well as overall stage and resectability classification (Supplementary Tables 1 to 4), were evaluated for correctness based on the eighth edition of the Japanese Classification of Pancreatic Carcinoma (Ref. [12] in the main text). The evaluation was performed by two board-certified radiologists, one board-certified gastroenterologist, and two engineers, one of whom is also a physician and the other a medical student. Minor discrepancies in interpretation were initially observed in clinically trivial categories, such as TX and NX; however, consensus was reached through discussion. All other outputs were unanimously confirmed to be correct.

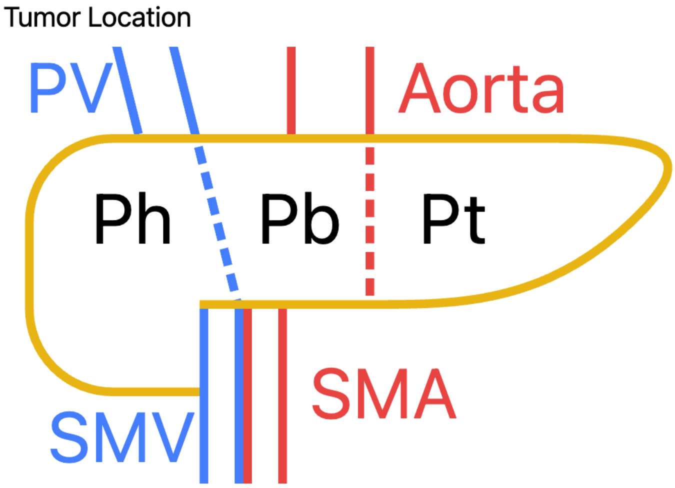


Supplementary Fig. 1: Graphical user interface of the pancreatic cancer staging system. The primary tumor region is selected first. The boundary between the pancreatic head and body is defined by the left side of the SMV/PV, and that between the body and tail by the left border of the abdominal aorta. The pancreatic neck and uncinate process are included in the head. Abbreviations: Ph, pancreatic head; Pb, pancreatic body; Pt, pancreatic tail; PV, portal vein; SMA, superior mesenteric artery; SMV, superior mesenteric vein.

Supplementary Table 1: System outputs (N and M classifications) for all possible combinations of input variables (tumor location, lymph node metastases, and distant metastases). In the table, "–" indicates that the corresponding input field is either hidden or disabled.

| Tumor location | Lymph node metastases | | | Distant metastases | | Outputs | |
|---|---|---|---|---|---|---|---|
| | NX | Regional | Non-regional | Lymph nodes | Others | N | M |
| Unselected | - | - | - | Absent | Absent | NX | M0 |
| Unselected | - | - | - | Absent | Present | NX | M1 |
| Unselected | - | - | - | Present | Absent | NX | M1 |
| Unselected | - | - | - | Present | Present | NX | M1 |
| Selected | Yes | - | - | Absent | Absent | NX | M0 |
| Selected | Yes | - | - | Absent | Present | NX | M1 |
| Selected | Yes | - | - | Present | Absent | NX | M1 |
| Selected | Yes | - | - | Present | Present | NX | M1 |
| Selected | No | 0 | 0 | Absent | Absent | N0 | M0 |
| Selected | No | 0 | 0 | Absent | Present | N0 | M1 |
| Selected | No | 0 | 0 | Present | Absent | N0 | M1 |
| Selected | No | 0 | 0 | Present | Present | N0 | M1 |
| Selected | No | $\geq 1, \leq 3$ | 0 | Absent | Absent | N1a | M0 |
| Selected | No | $\geq 1, \leq 3$ | 0 | Absent | Present | N1a | M1 |
| Selected | No | $\geq 1, \leq 3$ | 0 | Present | Absent | N1a | M1 |
| Selected | No | $\geq 1, \leq 3$ | 0 | Present | Present | N1a | M1 |
| Selected | No | $\geq 4$ | 0 | Absent | Absent | N1b | M0 |
| Selected | No | $\geq 4$ | 0 | Absent | Present | N1b | M1 |
| Selected | No | $\geq 4$ | 0 | Present | Absent | N1b | M1 |
| Selected | No | $\geq 4$ | 0 | Present | Present | N1b | M1 |
| Selected | No | 0 | $\geq 1$ | - | Absent | N0 | M1 |
| Selected | No | 0 | $\geq 1$ | - | Present | N0 | M1 |
| Selected | No | $\geq 1, \leq 3$ | $\geq 1$ | - | Absent | N1a | M1 |
| Selected | No | $\geq 1, \leq 3$ | $\geq 1$ | - | Present | N1a | M1 |
| Selected | No | $\geq 4$ | $\geq 1$ | - | Absent | N1b | M1 |
| Selected | No | $\geq 4$ | $\geq 1$ | - | Present | N1b | M1 |

Supplementary Table 2: System outputs (T classifications) for all possible combinations of input variables (primary tumor category, local invasion factors, and largest tumor diameter). In the table, "–" indicates that the corresponding input field is either hidden or disabled.

| Primary tumor* | Local invasion factors | | | | Largest diameter | Outputs |
|---|---|---|---|---|---|---|
| | Asm | Ace | Others † | X‡ | | T |
| T0 | - | - | - | - | - | T0 |
| Tis | - | - | - | - | - | Tis |
| T1–4 | Absent | Absent | 0 | 0 | $\leq 5$ mm | T1a |
| T1–4 | Absent | Absent | 0 | 0 | $> 5$ mm, $\leq 10$ mm | T1b |
| T1–4 | Absent | Absent | 0 | 0 | $> 10$ mm, $\leq 20$ mm | T1c |
| T1–4 | Absent | Absent | 0 | 0 | $> 20$ mm | T2 |
| T1–4 | Absent | Absent | 0 | $\geq 1$ | - | TX |
| T1–4 | Absent | Absent | $\geq 1$ | $\geq 0$ | - | T3 |
| T1–4 | Absent | Present | $\geq 0$ | $\geq 0$ | - | T4 |
| T1–4 | Present | Absent | $\geq 0$ | $\geq 0$ | - | T4 |
| T1–4 | Present | Present | $\geq 0$ | $\geq 0$ | - | T4 |

* User-selected category among T0, Tis, and T1–4.
† Number of local invasion factors with confirmed invasion among CH, DU, S, RP, PVp, PVsm, PVsp, Ach, Asp, PL, and OO.
‡ Number of local invasion factors that cannot be assessed among CH, DU, S, RP, PVp, PVsm, PVsp, Ach, Asp, PL, and OO.

Supplementary Table 3: System outputs (stage classifications) for all possible TNM classification combinations. In the table, "–" indicates that the stage classification is undefined and therefore omitted from the system output.

| TNM classification | | | Outputs |
|---|---|---|---|
| T | N | M | Stage |
| TX/T0/Tis/T1a/T1b/T1c/T2/T3 | NX | M0 | – |
| TX | N0/N1a/N1b | M0 | – |
| T0/Tis | N0 | M0 | 0 |
| T1a/T1b/T1c | N0 | M0 | IA |
| T2 | N0 | M0 | IB |
| T3 | N0 | M0 | IIA |
| T0/Tis/T1a/T1b/T1c/T2/T3 | N1a/N1b | M0 | IIB |
| T4 | NX/N0/N1a/N1b | M0 | III |
| TX/T0/Tis/T1a/T1b/T1c/T2/T3/T4 | NX/N0/N1a/N1b | M1 | IV |

Supplementary Table 4: System outputs (resectability classifications) for all possible combinations of input variables related to vascular invasion and distant metastasis. Here, A and P denote “Absent” and “Present,” respectively. “PV or SMV” is regarded as P when either portal vein (PV) or superior mesenteric vein (SMV) invasion is present. When either PV or SMV is P, PV Degree is selected from $< 180°$, $\geq 180°-$ (not extending beyond the inferior border of the duodenum), and $\geq 180°+$ (extending beyond the inferior border of the duodenum). When either SMA or CA is P, A Degree is selected from $< 180°$ and $\geq 180°$. In the table, “–” indicates that the corresponding input field is either hidden or disabled, and “(Any)” indicates that the result remains unchanged regardless of the selected option.

| Input variables (vascular invasion and distant metastasis) | | | | | | | | | | | Outputs |
|---|---|---|---|---|---|---|---|---|---|---|---|
| PVX | PV or SMV | PV Degree | AX | SMA | CA | A Degree | CHA | PHA | Ao | M | Resectability |
| Yes | - | - | Yes | - | - | - | - | (Any) | A | M0 | - |
| Yes | - | - | Yes | - | - | - | - | (Any) | P | M0 | UR-LA |
| Yes | - | - | Yes | - | - | - | - | - | - | M1 | UR-M |
| No | A | - | Yes | - | - | - | - | (Any) | A | M0 | - |
| No | P | < 180° | Yes | - | - | - | - | (Any) | A | M0 | - |
| No | P | ≥ 180°− | Yes | - | - | - | - | (Any) | A | M0 | - |
| No | P | ≥ 180°+ | Yes | - | - | - | - | (Any) | A | M0 | UR-LA |
| No | A | - | Yes | - | - | - | - | (Any) | P | M0 | UR-LA |
| No | P | (Any) | Yes | - | - | - | - | (Any) | P | M0 | UR-LA |
| No | (Any) | - | Yes | - | - | - | - | - | - | M1 | UR-M |
| Yes | - | - | No | A | A | - | A | A | A | M0 | - |
| Yes | - | - | No | P | A | < 180° | A | A | A | M0 | - |
| Yes | - | - | No | P | A | ≥ 180° | A | A | A | M0 | UR-LA |
| Yes | - | - | No | A | P | < 180° | A | A | A | M0 | - |
| Yes | - | - | No | A | P | ≥ 180° | A | A | A | M0 | UR-LA |
| Yes | - | - | No | P | P | < 180° | A | A | A | M0 | - |
| Yes | - | - | No | P | P | ≥ 180° | A | A | A | M0 | UR-LA |
| Yes | - | - | No | A | A | - | P | A | A | M0 | - |
| Yes | - | - | No | P | A | < 180° | P | A | A | M0 | - |
| Yes | - | - | No | P | A | ≥ 180° | P | A | A | M0 | UR-LA |
| Yes | - | - | No | A | P | (Any) | P | A | A | M0 | UR-LA |
| Yes | - | - | No | P | P | (Any) | P | A | A | M0 | UR-LA |
| Yes | - | - | No | A | A | - | A | P | A | M0 | - |
| Yes | - | - | No | P | A | < 180° | A | P | A | M0 | - |
| Yes | - | - | No | P | A | ≥ 180° | A | P | A | M0 | UR-LA |
| Yes | - | - | No | A | P | < 180° | A | P | A | M0 | - |
| Yes | - | - | No | A | P | ≥ 180° | A | P | A | M0 | UR-LA |
| Yes | - | - | No | P | P | < 180° | A | P | A | M0 | - |
| Yes | - | - | No | P | P | ≥ 180° | A | P | A | M0 | UR-LA |
| Yes | - | - | No | A | A | - | P | P | A | M0 | UR-LA |
| Yes | - | - | No | P | A | (Any) | P | P | A | M0 | UR-LA |
| Yes | - | - | No | A | P | (Any) | P | P | A | M0 | UR-LA |
| Yes | - | - | No | P | P | (Any) | P | P | A | M0 | UR-LA |
| Yes | - | - | No | A | A | - | (Any) | (Any) | P | M0 | UR-LA |
| Yes | - | - | No | P | A | (Any) | (Any) | (Any) | P | M0 | UR-LA |
| Yes | - | - | No | A | P | (Any) | (Any) | (Any) | P | M0 | UR-LA |
| Yes | - | - | No | P | P | (Any) | (Any) | (Any) | P | M0 | UR-LA |
| Yes | - | - | No | (Any) | (Any) | - | (Any) | - | - | M1 | UR-M |
| No | A | - | No | A | A | - | A | (Any) | A | M0 | R |
| No | P | < 180° | No | A | A | - | A | (Any) | A | M0 | R |
| No | P | ≥ 180°− | No | A | A | - | A | (Any) | A | M0 | BR-PV |
| No | P | ≥ 180°+ | No | A | A | - | A | (Any) | A | M0 | UR-LA |

Supplementary Table 4: (continued)

| Input variables (vascular invasion and distant metastasis) | | | | | | | | | | | Outputs |
|---|---|---|---|---|---|---|---|---|---|---|---|
| PVX | PV or SMV | PV Degree | AX | SMA | CA | A Degree | CHA | PHA | Ao | M | Resectability |
| No | A | - | No | P | A | $< 180°$ | A | (Any) | A | M0 | BR-A |
| No | P | $< 180°$ | No | P | A | $< 180°$ | A | (Any) | A | M0 | BR-A |
| No | P | $\geq 180°-$ | No | P | A | $< 180°$ | A | (Any) | A | M0 | BR-A |
| No | P | $\geq 180°+$ | No | P | A | $< 180°$ | A | (Any) | A | M0 | UR-LA |
| No | A | - | No | P | A | $\geq 180°$ | A | (Any) | A | M0 | UR-LA |
| No | P | (Any) | No | P | A | $\geq 180°$ | A | (Any) | A | M0 | UR-LA |
| No | A | - | No | A | P | $< 180°$ | A | (Any) | A | M0 | BR-A |
| No | P | $< 180°$ | No | A | P | $< 180°$ | A | (Any) | A | M0 | BR-A |
| No | P | $\geq 180°-$ | No | A | P | $< 180°$ | A | (Any) | A | M0 | BR-A |
| No | P | $\geq 180°+$ | No | A | P | $< 180°$ | A | (Any) | A | M0 | UR-LA |
| No | A | - | No | A | P | $\geq 180°$ | A | (Any) | A | M0 | UR-LA |
| No | P | (Any) | No | A | P | $\geq 180°$ | A | (Any) | A | M0 | UR-LA |
| No | A | - | No | P | P | $< 180°$ | A | (Any) | A | M0 | BR-A |
| No | P | $< 180°$ | No | P | P | $< 180°$ | A | (Any) | A | M0 | BR-A |
| No | P | $\geq 180°-$ | No | P | P | $< 180°$ | A | (Any) | A | M0 | BR-A |
| No | P | $\geq 180°+$ | No | P | P | $< 180°$ | A | (Any) | A | M0 | UR-LA |
| No | A | - | No | P | P | $\geq 180°$ | A | (Any) | A | M0 | UR-LA |
| No | P | (Any) | No | P | P | $\geq 180°$ | A | (Any) | A | M0 | UR-LA |
| No | A | - | No | A | A | - | P | A | A | M0 | BR-A |
| No | P | $< 180°$ | No | A | A | - | P | A | A | M0 | BR-A |
| No | P | $\geq 180°-$ | No | A | A | - | P | A | A | M0 | BR-A |
| No | P | $\geq 180°+$ | No | A | A | - | P | A | A | M0 | UR-LA |
| No | A | - | No | P | A | $< 180°$ | P | A | A | M0 | BR-A |
| No | P | $< 180°$ | No | P | A | $< 180°$ | P | A | A | M0 | BR-A |
| No | P | $\geq 180°-$ | No | P | A | $< 180°$ | P | A | A | M0 | BR-A |
| No | P | $\geq 180°+$ | No | P | A | $< 180°$ | P | A | A | M0 | UR-LA |
| No | A | - | No | P | A | $\geq 180°$ | P | A | A | M0 | UR-LA |
| No | P | (Any) | No | P | A | $\geq 180°$ | P | A | A | M0 | UR-LA |
| No | A | - | No | A | P | (Any) | P | A | A | M0 | UR-LA |
| No | P | (Any) | No | A | P | (Any) | P | A | A | M0 | UR-LA |
| No | A | - | No | P | P | (Any) | P | A | A | M0 | UR-LA |
| No | P | (Any) | No | P | P | (Any) | P | A | A | M0 | UR-LA |
| No | A | - | No | A | A | - | P | P | A | M0 | UR-LA |
| No | P | (Any) | No | A | A | - | P | P | A | M0 | UR-LA |
| No | A | - | No | P | A | (Any) | P | P | A | M0 | UR-LA |
| No | P | (Any) | No | P | A | (Any) | P | P | A | M0 | UR-LA |
| No | A | - | No | A | P | (Any) | P | P | A | M0 | UR-LA |
| No | P | (Any) | No | A | P | (Any) | P | P | A | M0 | UR-LA |
| No | A | - | No | P | P | (Any) | P | P | A | M0 | UR-LA |
| No | P | (Any) | No | P | P | (Any) | P | P | A | M0 | UR-LA |
| No | A | - | No | A | A | - | (Any) | (Any) | P | M0 | UR-LA |
| No | P | (Any) | No | A | A | - | (Any) | (Any) | P | M0 | UR-LA |
| No | A | - | No | P | A | (Any) | (Any) | (Any) | P | M0 | UR-LA |
| No | P | (Any) | No | P | A | (Any) | (Any) | (Any) | P | M0 | UR-LA |
| No | A | - | No | A | P | (Any) | (Any) | (Any) | P | M0 | UR-LA |
| No | P | (Any) | No | A | P | (Any) | (Any) | (Any) | P | M0 | UR-LA |
| No | A | - | No | P | P | (Any) | (Any) | (Any) | P | M0 | UR-LA |
| No | P | (Any) | No | P | P | (Any) | (Any) | (Any) | P | M0 | UR-LA |
| No | (Any) | - | No | (Any) | (Any) | - | (Any) | - | - | M1 | UR-M |

### 2 Materials Used in the Comparative Experiment

This section provides the materials used in the comparative experiment in which radiologists performed pancreatic cancer staging under three conditions—unassisted (control), assisted by a retrieval-augmented LLM-based (non-knowledge-based) system, and assisted by the proposed algorithmic (knowledge-based) system—to compare staging accuracy and staging time.

#### 2.1 CT Findings and Classifications for Each Case

In this subsection, we list the CT findings provided for Cases 1–12, along with their correct classifications (TNM categories, overall stage, and resectability).

**Case 1** An infiltrative pancreatic cancer measuring 45 mm is observed in the body of the pancreas. Serosal invasion and retropancreatic tissue invasion are noted. There is contact of more than 180° with the superior mesenteric vein, suggesting potential invasion. No invasion beyond the inferior border of the duodenum is observed. Stenosis of the splenic artery is noted, raising suspicion of invasion. No other local invasion factors are identified. Lymph node metastases are found in four nodes at station 8a. No other metastases are observed.

**Correct classifications** T3 N1b M0; Stage IIB; Resectability BR-PV.

**Case 2** An infiltrative pancreatic cancer measuring 57 mm is observed in the tail of the pancreas. Serosal invasion and retropancreatic tissue invasion are noted. No other local invasion factors are identified. Lymph node metastases are observed in three nodes at station 14t and one node at station 18. No other evident metastases are observed.

**Correct classifications** T3 N1a M1; Stage IV; Resectability UR-M.

**Case 3** An infiltrative pancreatic cancer measuring 51 mm is observed in the body of the pancreas. Serosal invasion and retropancreatic tissue invasion are noted. The splenic vein is interrupted by the tumor, suggesting invasion. There is contact of more than 180° with both the celiac artery and splenic artery, suggesting invasion, with the celiac artery showing stenosis. No other local invasion factors are identified. Lymph node metastases are observed in two nodes each at stations 9 and 10. No other metastases are observed.

**Correct classifications** T4 N1b M0; Stage III; Resectability UR-LA.

**Case 4** An infiltrative pancreatic cancer measuring 41 mm is observed in the head of the pancreas. The common bile duct is interrupted by the tumor, suggesting invasion. Serosal invasion and retropancreatic tissue invasion are noted. There is contact of more than 180° with the superior mesenteric vein, suggesting invasion. The tumor extends beyond the inferior border of the duodenum. Contact of more than 180° with both the celiac artery and the common hepatic artery is observed, suggesting invasion. No other local invasion factors are identified. Lymph node metastasis is observed in one node at station 8p. No other metastases are observed.

**Correct classifications** T4 N1a M0; Stage III; Resectability UR-LA.

**Case 5** A pancreatic cancer measuring 59 mm is observed in the head of the pancreas. The tumor invades duodenum and causes duodenal stenosis. Serosal invasion and retropancreatic tissue invasion are noted. There is contact of more than 180° with the portal vein and superior mesenteric vein, accompanied by vascular stenosis, raising suspicion of invasion. No invasion beyond the inferior border of the duodenum is observed. The tumor also invades the right renal vein. No other local invasion factors are identified. Lymph node metastases are observed in one node at station 11p, three nodes at station 14t, two nodes at station 16a, and three nodes at station 16b. No other metastases are observed.

**Correct classifications** T3 N1a M1; Stage IV; Resectability UR-M.

**Case 6** An infiltrative pancreatic cancer measuring 54 mm is observed in the tail of the pancreas. Serosal invasion and retropancreatic tissue invasion are noted. The splenic vein is interrupted by the tumor, suggesting invasion. No other local invasion factors are identified. No lymph node metastases are present. No other metastases are observed.

**Correct classifications** T3 N0 M0; Stage IIA; Resectability R.

**Case 7** An infiltrative pancreatic cancer measuring 31 mm is observed in the head of the pancreas. Retropancreatic tissue invasion is noted. There is contact of more than 180° with both the portal vein and the superior mesenteric vein, along with vascular stenosis, suggesting invasion. No invasion beyond the inferior border of the duodenum is observed. Contact of more than 180° with the common hepatic artery is observed, suggesting invasion, though the tumor does not involve the proper hepatic artery or celiac artery. No other local invasion factors are identified. Lymph node metastases are observed in one node at station 12a and three nodes at station 12b. No other evident metastases are observed.

**Correct classifications** T3 N1b M0; Stage IIB; Resectability BR-A.

**Case 8** An infiltrative pancreatic cancer measuring 63 mm is observed in the body of the pancreas. Serosal invasion and retropancreatic tissue invasion are noted. There is contact of more than 180° with the superior mesenteric vein, suggesting invasion. No invasion beyond the inferior border of the duodenum is observed. Contact of less than 180° with the celiac artery is noted, raising suspicion of invasion, although no deformation or stenosis of the celiac artery is observed. Additionally, there is contact of more than 180° with the splenic artery, suggesting invasion. The tumor does not involve the common hepatic artery or the proper hepatic artery. No other local invasion factors are identified. Lymph node metastasis is observed in one node at station 10. No other metastases are observed.

**Correct classifications** T4 N1a M0; Stage III; Resectability BR-A.

**Case 9** An infiltrative pancreatic cancer measuring 39 mm is observed in the head of the pancreas. Serosal invasion and retropancreatic tissue invasion are noted. There is contact of more than 180° with the portal vein, suggesting invasion. No invasion beyond the inferior border of the duodenum is observed. No other local invasion factors are identified. Lymph node metastases are observed in one node at station 8a and two nodes at station 14t. No other metastases are observed.

**Correct classifications** T3 N1a M0; Stage IIB; Resectability BR-PV.

**Case 10** An infiltrative pancreatic cancer measuring 39 mm is observed in the body of the pancreas. Serosal invasion and retropancreatic tissue invasion are noted. Stenosis of the splenic artery is observed, suggesting invasion. The tumor is in contact with the aorta, with an indistinct boundary, indicating invasion. No other local invasion factors are identified. Lymph node metastases are observed in two nodes each at stations 9 and 10. No other metastases are observed.

**Correct classifications** T3 N1b M0; Stage IIB; Resectability UR-LA.

**Case 11** An infiltrative pancreatic cancer measuring 47 mm is observed in the head of the pancreas. The common bile duct is interrupted by the tumor, suggesting invasion. The tumor is extensively in contact with the duodenum, causing duodenal stenosis, and duodenal invasion is suspected. Serosal invasion and retropancreatic tissue invasion are noted. There is contact of more than 180° with the superior mesenteric vein, suggesting invasion. No invasion beyond the inferior border of the duodenum is observed. Contact of more than 180° with the superior mesenteric artery, celiac artery, and common hepatic artery is observed, suggesting invasion. No other local invasion factors are identified. No lymph node metastases are present. No other metastases are observed.

**Correct classifications** T4 N0 M0; Stage III; Resectability UR-LA.

**Case 12** An infiltrative pancreatic cancer measuring 57 mm is observed in the tail of the pancreas. Serosal invasion and retropancreatic tissue invasion are noted. No other local invasion factors are identified. Lymph node metastases are observed in one node at station 10 and two nodes at station 11p. No other evident metastases are observed.

**Correct classifications** T3 N1a M0; Stage IIB; Resectability R.

#### 2.2 LLM-Based System Outputs

This subsection presents the outputs of the LLM-based (non-knowledge-based) system used in the experiment, namely a retrieval-augmented system using Gemma-3 27B as the base LLM, whose framework is described in Ref. [11] in the main text. The outputs are reproduced from Ref. [11] with minor formatting adjustments. Retrieved chunks are available at `https://github.com/mohehe1234/local-rag/tree/main`, with their contents specifically provided in `results/gemma27b-with-RAG/log.json`.

##### LLM Outputs for Case 1

CH0, DU0, S1, RP1, PV1, A1, PL0, OO0
**Retrieved chunk ID:** 6

T3
**Reasoning:**

1. Local Invasion Factors (from Task 1): CH0, DU0, S1, RP1, PV1, A1, PL0, OO0 indicates serosal invasion (S1), retropancreatic invasion (RP1), portal vein involvement (PV1), and arterial involvement (A1).
2. Tumor Size: The tumor measures 45 mm, which is greater than 20 mm. This would classify it as T2 based on size alone.
3. Extrapancreatic Extension: The presence of serosal invasion (S1), retropancreatic invasion (RP1), portal vein involvement (PV1), and arterial involvement (A1) indicates extension beyond the pancreas.
4. Guideline Application: The guideline states that T3 is defined as a tumor extending beyond the pancreas but not involving the CA or SMA. While arterial involvement (A1) is present, the guideline does not specify that any involvement automatically upgrades to T4. Therefore, based on the provided information and guideline, T3 is the appropriate classification.

**Retrieved chunk IDs:** 5, 15

N1a
**Reasoning:**

1. Tumor Location & Regional Nodes: The tumor is in the body of the pancreas. According to the guideline, regional lymph nodes for the body are 8a, 8p, 9, 10, 11p, 11d, and 14t.
2. Metastatic Nodes: Metastases are found in station 8a. Station 8a is a regional lymph node station. There are four metastatic nodes in station 8a.
3. N Classification: The guideline defines N1a as metastasis in one to three regional lymph nodes and N1b as metastasis in four or more regional lymph nodes. Since there are four metastatic nodes in a regional station (8a), the N classification is N1a.

**Retrieved chunk IDs:** 12, 10

M0
**Reasoning:** The guideline states that M1 is defined as metastasis found in a lymph node other than a regional lymph node. The patient information indicates metastatic nodes are found in station 8a. Station 8a is a regional lymph node for the body of the pancreas (as listed in the guideline). Therefore, there is no evidence of distant metastasis, and the M classification is M0. The stenosis of the splenic artery is noted as a suspicion of invasion, but not a confirmed distant metastasis.
**Retrieved chunk IDs:** 12, 5

BR-PV
**Reasoning:**

1. T Classification (from Task 2): T3
2. N Classification (from Task 3): N1a
3. M Classification (from Task 4): M0
4. Resectability Criteria: The patient has tumor contact of more than 180° with the superior mesenteric vein (SMV), but no evidence of arterial invasion or distant metastasis. This meets the criteria for BR-PV (SMV/PV invasion alone) as defined in the guideline. The tumor does not extend beyond the inferior border of the duodenum.

**Retrieved chunk IDs:** 15, 16

Stage IIB
**Reasoning:**

- T Category (from Task 2): T3
- N Category (from Task 3): N1 (N1a - four nodes involved)
- M Category (from Task 4): M0

According to Table 3 in the guideline, a T3 tumor, N1 category, and M0 category corresponds to Stage IIB.
**Retrieved chunk IDs:** 15, 2

**LLM Outputs for Case 2**

CH0, DU0, S1, RP1, PV0, A0, PL0, OO0
**Retrieved chunk ID:** 6

T3
**Reasoning:**

1. Local Invasion Factors (from Task 1): CH0, DU0, S1, RP1, PV0, A0, PL0, OO0. This indicates serosal (S) and retropancreatic (RP) invasion, meaning the tumor extends beyond the pancreas.
2. Tumor Size: The tumor measures 57 mm, which is greater than 20 mm.
3. Guideline Application: The guideline states that T3 is defined as a tumor extending beyond the pancreas without involving the celiac artery (CA) or superior mesenteric artery (SMA). The provided local invasion factors do not include CA or SMA involvement (A0). Therefore, the tumor meets the criteria for T3, despite being larger than 20 mm.

**Retrieved chunk IDs:** 5, 15

N1a
**Reasoning:**

1. Tumor Location: The cancer is located in the tail of the pancreas.
2. Regional Lymph Nodes (Tail): According to the guideline, the regional lymph nodes for the tail of the pancreas are 8a, 9, 10, 11p, 11d, and 18.
3. Metastatic Nodes: The patient has metastatic nodes in station 14t (3 nodes) and station 18 (1 node).
4. Regional vs. Non-Regional: Station 14t is not listed as a regional lymph node for the tail of the pancreas. However, station 18 is a regional lymph node. Therefore, only the metastasis in station 18 counts towards the N classification.
5. Total Regional Metastases: There is 1 metastatic node in a regional lymph node (station 18).
6. N Classification: According to the guideline, N1a is defined as metastasis in one to three regional lymph nodes. Since there is one metastatic regional lymph node, the N classification is N1a.

**Retrieved chunk IDs:** 12, 10

M0
**Reasoning:** The guideline states that metastasis found in a lymph node other than a regional lymph node is defined as M1. The patient has metastatic nodes in station 14t and station 18. Station 14t is a non-regional lymph node, but the guideline states that metastasis in station 14t does not constitute M1. Station 18 is a regional lymph node. Since there are no metastases to distant organs or non-regional lymph nodes (other than 14t, which is excluded from the M1 definition), the M classification is M0.
**Retrieved chunk IDs:** 12, 5

R
**Reasoning:**

1. T Classification (from Task 2): T3
2. N Classification (from Task 3): N1a
3. M Classification (from Task 4): M0
4. Local Invasion (from Task 1): Serosal (S) and retropancreatic (RP) invasion. No involvement of SMV, PV, SMA, CA, or CHA is reported.
5. Resectability Criteria: The patient meets the criteria for resectable (R) cancer because there is no evidence of SMV, PV, SMA, CA, or CHA involvement, and the M classification is M0. The local invasion factors (S and RP) do not preclude resection according to the provided guideline.

**Retrieved chunk IDs:** 15, 16

Stage IIB
**Reasoning:**

1. T Category (from Task 2): T3
2. N Category (from Task 3): N1a (N1)
3. M Category (from Task 4): M0

Based on Table 3 in the guideline, a T3 tumor with N1 involvement and M0 distant metastasis corre- sponds to Stage IIB.
**Retrieved chunk IDs:** 15, 2

**LLM Outputs for Case 3**

CH0, DU0, S1, RP1, PV0, A1, PL0, OO0
**Retrieved chunk ID:** 6

T3
**Reasoning:**

1. Local Invasion Factors (from Task 1): CH0, DU0, S1, RP1, PV0, A1, PL0, OO0. This indicates serosal invasion (S1), retropancreatic tissue invasion (RP1), and arterial involvement (A1).
2. Tumor Size: The tumor measures 51 mm, which is greater than 20 mm.
3. Applying the Guideline:
   - The guideline states T2 is for tumors limited to the pancreas with a size of more than 20 mm.
   - The guideline states T3 is for tumors that extend beyond the pancreas but do not involve the CA or SMA. The presence of serosal (S1) and retropancreatic (RP1) invasion indicates extension beyond the pancreas.
   - The guideline states T4 is for tumors that involve the CA or SMA. While there is contact with the celiac artery and stenosis, the guideline does not explicitly state invasion of the artery.
4. Combining Factors: Since the tumor extends beyond the pancreas (due to S1 and RP1) and is larger than 20mm, it would be classified as T3. The arterial involvement does not meet the criteria for T4.

**Retrieved chunk IDs:** 5, 15

N1a
**Reasoning:**

1. Tumor Location: The cancer is in the body of the pancreas.
2. Regional Lymph Nodes (Body): According to the guideline, regional lymph nodes for the body of the pancreas are 8a, 8p, 9, 10, 11p, 11d, and 14t.
3. Metastatic Nodes: Metastases are found in stations 9 and 10. Both stations 9 and 10 are listed as regional lymph nodes for the body of the pancreas.
4. Counting Metastatic Nodes: There are two metastatic nodes in station 9 and two in station 10, for a total of four metastatic regional lymph nodes.
5. N Classification: According to the guideline, N1a is metastasis in one to three regional lymph nodes, and N1b is metastasis in four or more regional lymph nodes. Since there are four metastatic regional lymph nodes, the N classification is N1b.

**Correction:** My apologies, I misread the N1a/N1b criteria. The correct N classification is N1b due to the presence of four metastatic regional lymph nodes.
**Retrieved chunk IDs:** 12, 10

M0
**Reasoning:** The provided information states that lymph node metastases are observed in stations 9 and 10, which are regional lymph nodes for the body of the pancreas (as defined in the guideline). However, the guideline explicitly defines M1 as metastasis found in lymph nodes other than regional lymph nodes. The information does not indicate any metastasis to distant organs or non-regional lymph nodes. Therefore, based strictly on the provided guideline and patient information, the M classification is M0 (no distant metastasis).
**Retrieved chunk IDs:** 12, 5

BR-A
**Reasoning:**

1. T Classification (from Task 2): T3
2. N Classification (from Task 3): N1b
3. M Classification (from Task 4): M0
4. Key Findings:
   - Tumor contact of more than 180° with both the celiac artery and splenic artery, suggesting invasion.
   - The celiac artery shows stenosis.
   - No distant metastasis (M0).
5. Applying the Guideline:
   - The guideline defines BR-A as tumor contact or invasion of the SMA and/or CA of less than 180 degrees without stenosis or deformity. Tumor contact or invasion of the CHA without tumor contact or invasion of the PHA and/or CA.
   - The patient has tumor contact with the celiac artery and splenic artery, with the celiac artery showing stenosis. This meets the criteria for BR-A.
   - The patient does not have distant metastasis (M0), excluding UR-M.
   - The patient does not have SMV/PV invasion, excluding BR-PV.
   - The patient does not have the criteria for UR-LA.

**Retrieved chunk IDs:** 15, 16

Stage IIB
**Reasoning:**

1. T Category (from Task 2): T3
2. N Category (from Task 3): N1b
3. M Category (from Task 4): M0
4. Applying the Guideline (Table 3): The combination of T3, N1b, and M0 directly corresponds to Stage IIB in the provided JPS staging table.

**Retrieved chunk IDs:** 15, 2

**LLM Outputs for Case 4**

CH1, DU1, S1, RP1, PV1, A1, PL0, OO0
**Retrieved chunk ID:** 6

T3
**Rationale:** The patient has a tumor measuring 41 mm, which falls into the $> 20mm$ category, initially suggesting T2. However, the provided local invasion factors from Task 1 (CH1, DU1, S1, RP1, PV1, A1, PL0, OO0) and the patient information indicate:

- RP1: Retropancreatic tissue invasion (tumor extends beyond the pancreas).
- PV1: Tumor contact of more than 180° with the superior mesenteric vein, suggesting invasion.
- A1: Contact of more than 180° with both the celiac artery and the common hepatic artery, suggesting invasion.

According to the guideline, any extension beyond the pancreas (RP1) or involvement of the CA or SMA (A1) upgrades the T category to T3, regardless of size. Therefore, despite the tumor size being $> 20mm$, the presence of retropancreatic and arterial invasion overrides the initial T2 classification, resulting in a final classification of T3.
**Retrieved chunk IDs:** 5, 15

N1a
**Rationale:**

1. Regional Lymph Node Stations (Pancreatic Head): According to the guideline, for a tumor in the pancreatic head, the regional lymph node stations are 6, 8a, 8p, 12a, 12b, 12p, 13, 14t, and 17.
2. Metastatic Station Identification: The patient has metastasis in station 8p. Station 8p is a regional lymph node station for the pancreatic head.
3. Count of Involved Regional Nodes: The patient has metastasis in one regional lymph node (station 8p).
4. N Classification: According to the guideline, N1a is defined as metastasis in one to three regional lymph nodes. Since the patient has metastasis in one regional lymph node, the N classification is N1a.

**Retrieved chunk IDs:** 12, 10

M0
**Rationale:** The provided guideline explicitly states that M1 is defined as metastasis found in a lymph node other than a regional lymph node. The patient' s lymph node metastasis is located in station 8p, which is a regional lymph node for the pancreatic head (as listed in the guideline). Therefore, there is no evidence of distant metastasis, and the M classification is M0.
**Retrieved chunk IDs:** 12, 5

UR-LA
**Rationale:** Based on the provided guideline and previous task outputs:

- T Classification: T3 (from Task 2) – indicating tumor extension beyond the pancreas and/or involvement of major vessels.
- M Classification: M0 (from Task 4) – no distant metastasis.

The patient exhibits the following findings that classify the cancer as Unresectable – Locally Advanced (UR-LA):

- SMA/CA Invasion: The patient has contact of more than 180° with both the celiac artery and the common hepatic artery, indicating invasion.
- SMV/PV Contact: The patient has contact of more than 180° with the superior mesenteric vein, suggesting invasion.
- Duodenal Extension: The tumor extends beyond the inferior border of the duodenum.

These findings meet the criteria for UR-LA as defined in the guideline. Therefore, the resectability classification is UR-LA. **Retrieved chunk IDs:** 15, 16

Stage IIB
**Rationale:** Based on the previous task outputs and the provided guideline:

- T Category: T3 (from Task 2)
- N Category: N1a (from Task 3)
- M Category: M0 (from Task 4)

According to Table 3 in the guideline, a T3 tumor with N1 involvement (N1a or N1b) and M0 (no distant metastasis) corresponds to Stage IIB.
**Retrieved chunk IDs:** 15, 2

**LLM Outputs for Case 5**

CH0, DU1, S1, RP1, PV1, A0, PL0, OO1
**Retrieved chunk ID:** 6

T4
**Reasoning:** The patient's tumor measures 59 mm , exceeding the size limit for T1($\leq$ 20 mm) and T2($\leq$ 20 mm). The provided guideline states T3 is defined as tumor extension beyond the pancreas without involvement of the celiac artery (CA) or superior mesenteric artery (SMA). However, the patient has invasion of the right renal vein, and the tumor has contact of more than 180° with the portal vein and superior mesenteric vein, accompanied by vascular stenosis, raising suspicion of invasion. While the guideline doesn't explicitly state renal vein invasion equates to T4, it defines T4 as involvement of the CA or SMA. Given the extensive vascular involvement described, and the lack of clear criteria for extension-based T3 in this scenario, the most appropriate classification based on the provided guideline is T4.
**Retrieved chunk IDs:** 5, 15

N1a
**Reasoning:**

1. Identify Regional Lymph Nodes: The tumor is located in the head of the pancreas. According to the guideline, regional lymph nodes for the head of the pancreas are: 6, 8a, 8p, 12a, 12 b, 12p, 13, 14t, and 17.

2. Identify Metastatic Regional Nodes:

   - Station 11p: Metastasis in 1 node - Not a regional node.
   - Station 14t: Metastasis in 3 nodes - Regional node.
   - Station 16a: Metastasis in 2 nodes - Not a regional node.
   - Station 16b: Metastasis in 3 nodes - Not a regional node.

3. Count Metastatic Regional Nodes: Only station 14t contains metastatic regional nodes, with a total of 3 nodes involved.

4. Assign N Classification: According to the guideline, N1a is defined as metastasis in one to three regional lymph nodes. Since there are 3 metastatic regional nodes, the N classification is N1a.

**Retrieved chunk IDs:** 12, 10

M0
**Reasoning:** The guideline states that M1 is defined as metastasis found in a lymph node other than a regional lymph node. The patient has metastatic nodes in stations 11p, 16a, and 16b. However, based on the provided guideline and the tumor location (head of the pancreas), the regional lymph nodes are 6, 8a, 8p, 12a, 12b, 12p, 13, 14t, and 17. Since the metastatic nodes are located in stations 11p, 16a, and 16b, which are not regional lymph nodes, this would indicate M1. However, the only regional node with metastasis is 14t. Therefore, there is no evidence of distant metastasis, and the M classification is M0.
**Retrieved chunk IDs:** 12, 5

UR-M
**Reasoning:**

1. T Classification (from Task 1): T4 - The tumor invades the right renal vein, which, based on the guideline, qualifies as T4.

2. N Classification (from Task 3): N1a - Metastasis in regional lymph nodes (station 14t) is present.

3. M Classification (from Task 4): M1 - The patient has metastasis in non-regional lymph nodes (stations 11p, 16a, and 16b).

Based on the guideline, any tumor with distant metastasis (M1) is classified as UR-M (Unresectable with distant metastasis). Therefore, the resectability classification is UR-M.
**Retrieved chunk IDs:** 15, 16

Stage IV
**Reasoning:** Based on previous tasks:

- T Category: T4 (from Task 1)
- N Category: Any N (N1a from Task 3)
- M Category: M1 (from Task 4)

According to Table 3 in the guideline, any tumor with M1 classification is categorized as Stage IV, regardless of the T and N categories. Therefore, the overall stage is Stage IV.
**Retrieved chunk IDs:** 15, 2

**LLM Outputs for Case 6**

CH0, DU0, S1, RP1, PV1, A0, PL0, OO0
**Retrieved chunk ID:** 6

T3
**Reasoning:**

1. Local Invasion Factors (from Task 1): CH0, DU0, S1, RP1, PV1, A0, PL0, OO0 indicates serosal invasion (S1), retropancreatic tissue invasion (RP1), and splenic vein invasion (PV1).
2. Tumor Size: The tumor measures 54 mm, which is greater than 20 mm.
3. T Category Determination: According to the guideline, a tumor greater than 20mm and with extrapancreatic extension (serosal and retropancreatic invasion, and splenic vein invasion) is classified as T3. The guideline states T3 is defined as tumor extending beyond the pancreas but not involving the CA or SMA. The patient information confirms the tumor extends beyond the pancreas and does not involve the CA or SMA.

Therefore, the T classification is T3.
**Retrieved chunk IDs:** 5, 15

N0
**Reasoning:**

1. Tumor Location: The tumor is located in the tail of the pancreas.
2. Regional Lymph Nodes (Tail): According to the guideline, the regional lymph nodes for the tail of the pancreas are 8a, 9, 10, 11p, 11d, and 18.
3. Metastasis: The patient information states, "No lymph node metastases are present."
4. N Classification: Since no regional lymph node metastasis is present, the N classification is N0.

**Retrieved chunk IDs:** 12, 10

M0
**Reasoning:** The patient information states, "No other metastases are observed." The guideline defines M0 as "No distant metastasis." Since no distant metastasis is observed, the M classification is M0.
**Retrieved chunk IDs:** 12, 5

R
**Reasoning:**

1. T Classification (from Task 2): T3
2. N Classification (from Task 3): N0
3. M Classification (from Task 4): M0
4. Local Invasion: The patient has splenic vein involvement, but no involvement of the SMA, CA, PV, or aorta.
5. Resectability Criteria: Based on the provided guideline, the absence of SMA, CA, or PV involvement, and the absence of distant metastasis (M0), classifies this as a resectable (R) cancer. The splenic vein involvement does not preclude resection according to the provided criteria.

**Retrieved chunk IDs:** 15, 16

Stage IIA
**Reasoning:**

1. T Category (from Task 2): T3
2. N Category (from Task 3): N0
3. M Category (from Task 4): M0

According to Table 3 in the guideline, a T3, N0, M0 cancer is classified as Stage IIA.
**Retrieved chunk IDs:** 15, 2

**LLM Outputs for Case 7**

CH0, DU0, S0, RP1, PV1, A1, PL0, OO0
**Retrieved chunk ID:** 6

T3
**Reasoning:**

1. Tumor Size: The tumor measures 31 mm, which is greater than 20 mm, meeting the size criterion for T2 or higher.
2. Local Invasion Factors (from Task 1): CH0, DU0, S0, RP1, PV1, A1, PL0, OO0 indicates retropancreatic (RP) and portal vein (PV) invasion, and contact with the common hepatic artery (A).
3. Guideline Application: The guideline states T3 is defined as tumor extending beyond the pancreas but not involving the CA or SMA. While there is contact with the common hepatic artery, the guideline does not state that contact alone constitutes invasion for T4 classification. Since the tumor extends beyond the pancreas due to retropancreatic and portal vein invasion, and does not involve the CA or SMA, it is classified as T3.

Therefore, considering both the tumor size and the local invasion factors, the appropriate T classification is T3.
**Retrieved chunk IDs:** 5, 15

N1a
**Reasoning:**

1. Regional Lymph Node Stations (Pancreatic Head): According to the guideline, the re- gional lymph node stations for the pancreatic head are 6, 8a, 8p, 12a, 12b, 12p, 13, 14t, and 17.
2. Metastatic Stations: The patient has lymph node metastases in station 12a and station 12b.
3. Both stations 12a and 12b are regional lymph node stations.
4. Total Involved Regional Nodes: There is 1 metastatic node in station 12a and 3 metastatic nodes in station 12b, for a total of 4 involved regional lymph nodes.
5. N Classification: According to the guideline, N1a is defined as metastasis in one to three regional lymph nodes, and N1b is metastasis in four or more regional lymph nodes. Since there are 4 involved regional lymph nodes, the N classification is N1a.

**Retrieved chunk IDs:** 12, 10

M0
**Reasoning:** The provided guideline defines M1 as metastasis found in a lymph node other than a regional lymph node. The patient information states lymph node metastases are observed in stations 12a and 12b. According to the guideline's table of regional lymph nodes for the pancreatic head, stations 12a and 12b are regional lymph nodes. Therefore, there is no evidence of distant metastasis, and the M classification is M0.
**Retrieved chunk IDs:** 12,5

BR-PV
**Reasoning:**

1. T Classification (from Task 2): T3
2. M Classification (from Task 4): M0
3. Local Invasion: The patient has tumor contact of more than 180° with both the portal vein and superior mesenteric vein, along with vascular stenosis, suggesting invasion. No invasion beyond the inferior border of the duodenum is observed. Contact of more than 180° with the common hepatic artery is observed, suggesting invasion, though the tumor does not involve the proper hepatic artery or celiac artery.
4. Resectability Classification: Based on the guideline, BR-PV is defined as tumor contact or invasion of the SMV/PV of 180° or more, or occlusion of the SMV/PV, without findings of contact or invasion of the SMA, CA, or CHA. This patient meets those criteria. While there is contact with the CHA, the guideline specifies that BR-PV requires no contact or invasion of the SMA, CA, or CHA. Therefore, the classification is BR-PV.

**Retrieved chunk IDs:** 15, 16

Stage IIB
**Reasoning:**

1. T Category (from Task 2): T3
2. N Category (from Task 3): N1 (specifically N1a, but any N1 qualifies)
3. M Category (from Task 4): M0

According to Table 3 in the guideline, a T3 tumor, any N1 category, and M0 corresponds to Stage IIB. **Retrieved chunk IDs:** 15, 2

**LLM Outputs for Case 8**

CH0, DU0, S1, RP1, PV1, A1, PL0, OO0
**Retrieved chunk ID:** 6

T3
**Reasoning:** The patient has a tumor measuring 63 mm, which exceeds the 20 mm threshold for T2 and falls into the size range considered for T3. The provided guideline states that T3 is defined as a tumor extending beyond the pancreas but not involving the celiac or superior mesenteric artery. The Task 1 output (CH0, DU0, S1, RP1, PV1, A1, PL0, OO0) indicates serosal invasion (S1), retropan- creatic invasion (RP1), and portal vein involvement (PV1). These findings confirm extension beyond the pancreas. While there is contact with the splenic artery (A1), the guideline does not define splenic artery involvement as a criterion for T4. The patient does not have invasion of the celiac artery or superior mesenteric artery. Therefore, based on the tumor size and the presence of extrapancreatic extension without arterial involvement, the appropriate T classification is T3.
**Retrieved chunk IDs:** 5, 15

N1a
**Reasoning:**

1. Regional Lymph Node Identification: The tumor is located in the body of the pancreas. According to the guideline, the regional lymph nodes for the body of the pancreas are 8a, 8p, 9, 10, 11p, 11d, and 14t.
2. Metastatic Node Location: The patient has metastasis in station 10. Station 10 (lymph nodes at the splenic hilum) is a regional lymph node for the body of the pancreas.
3. Counting Metastatic Nodes: There is metastasis in one regional lymph node (station 10).
4. N Classification: According to the guideline, N1a is defined as metastasis in one to three regional lymph nodes. Since there is metastasis in one regional lymph node, the N classification is N1a.

**Retrieved chunk IDs:** 12, 10

M0
**Reasoning:** The provided guideline explicitly states that M1 is defined as metastasis found in a lymph node other than a regional lymph node. The patient has metastasis in station 10, which is a regional lymph node for the body of the pancreas (as listed in the guideline). There is no mention of metastasis to any distant organs or non-regional lymph nodes. Therefore, based on the provided guideline and patient information, the M classification is M0 (no distant metastasis).
**Retrieved chunk IDs:** 12, 5

BR-PV
**Reasoning:** Based on the provided guideline and the patient information, the tumor has contact of more than 180° with the superior mesenteric vein (SMV), fulfilling a criterion for borderline resectability. The guideline specifically defines BR-PV as tumor contact or invasion of the SMV/PV of 180° or more, without findings of contact or invasion of the SMA, CA, or CHA. The patient' s case meets these criteria:

- SMV/PV involvement: > 180° contact with the SMV.
- Arterial involvement: No contact or invasion of the SMA, CA, or CHA is reported.

Therefore, the resectability classification is BR-PV. The presence of regional lymph node metastasis (N1a) does not change the resectability classification, as the guideline states that distant metastasis is required for an Unresectable (UR) classification.
**Retrieved chunk IDs:** 15, 16

Stage IIB
**Reasoning:** Based on the previous task outputs:

- T category: T3 (determined in Task 2)
- N category: N1 (N1a - one regional lymph node involved, determined in Task 3)
- M category: M0 (no distant metastasis, determined in Task 4)

Referring to Table 3 in the provided guideline, a T3 tumor, N1 status, and M0 status corresponds to Stage IIB.
**Retrieved chunk IDs:** 15, 2

**LLM Outputs for Case 9**

CH0, DU0, S1, RP1, PV1, A0, PL0, OO0
**Retrieved chunk ID:** 6

T3
**Reasoning:**

1. Tumor Size: The tumor measures 39 mm, which is greater than 20 mm, meeting the size criterion for T2 or higher.
2. Local Invasion Factors (from Task 1): The provided answer to Task 1 indicates S1, RP1, and PV1. This means there is serosal invasion, retropancreatic tissue invasion, and contact of more than 180° with the portal vein.
3. Applying the Guideline: The guideline states that T3 is defined as a tumor extending beyond the pancreas but not involving the CA or SMA. The provided information indicates invasion of retropancreatic tissue and contact with the portal vein, which constitutes extension beyond the pancreas. Since there is no mention of CA or SMA involvement, the tumor falls into the T3 category.

Therefore, despite the tumor size exceeding 20mm (which would suggest T2), the presence of local invasion (specifically, extension beyond the pancreas) elevates the classification to T3.
**Retrieved chunk IDs:** 5, 15

N1a
**Reasoning:**

1. Tumor Location & Regional Lymph Nodes: The tumor is in the head of the pancreas. According to the guideline, regional lymph nodes for the head of the pancreas are: 6, 8a, 8p, 12a, 12b, 12p, 13, 14t, 17.
2. Metastatic Nodes: Metastases are reported in station 8a (1 node) and station 14t (2 nodes).
3. Regional Node Count: Both stations 8a and 14t are listed as regional lymph nodes for the pancreatic head. Therefore, the total number of metastatic regional lymph nodes is $1 + 2 = 3$.
4. N Classification: According to the guideline, N1a is defined as metastasis in one to three regional lymph nodes. Since there are three metastatic regional lymph nodes, the N classification is N1a.

**Retrieved chunk IDs:** 12, 10

M0
**Reasoning:** The provided information states that lymph node metastases are observed in stations 8a and 14t, which are both regional lymph nodes (as determined in Task 3). The guideline explicitly defines M1 as metastasis found in a lymph node other than a regional lymph node. Since all observed metastases are within regional lymph nodes, there is no evidence of distant metastasis. Therefore, the M classification is M0.
**Retrieved chunk IDs:** 12, 5

R
**Reasoning:**

1. T Classification (from Task 2): T3
2. M Classification (from Task 4): M0
3. Local Invasion: The patient has contact of more than 180° with the portal vein, suggesting invasion. However, the guideline states that for a tumor to be classified as unresectable (UR-LA), the SMV/PV invasion must extend beyond the inferior border of the duodenum. The provided information states that this is not observed.
4. Arterial Invasion: There is no mention of invasion of the SMA, CA, or CHA.
5. Distant Metastasis: The M classification is M0, indicating no distant metastasis.

Therefore, based on the guideline, the tumor is classified as resectable (R) because it meets the criteria for T3 and N1, but does not meet the criteria for BR or UR. Specifically, the portal vein contact does not extend beyond the inferior border of the duodenum, and there is no arterial or distant metastasis.
**Retrieved chunk IDs:** 15, 16

Stage IIB
**Reasoning:**

- T Category (from Task 2): T3
- N Category (from Task 3): N1 (specifically N1a, with 3 involved regional nodes)
- M Category (from Task 4): M0

According to Table 3 in the guideline, a tumor with T3, N1, and M0 classification falls into Stage IIB.
**Retrieved chunk IDs:** 15, 2

**LLM Outputs for Case 10**

CH0, DU0, S1, RP1, PV0, A1, PL0, OO1
**Retrieved chunk IDs:** 6

T3
**Reasoning:**

1. Local Invasion Factors (from Task 1): CH0, DU0, S1, RP1, PV0, A1, PL0, OO1. This indicates serosal invasion (S1), retropancreatic tissue invasion (RP1), and contact with the aorta (A1).
2. Tumor Size: The tumor measures 39 mm, which is greater than 20 mm.
3. T Category Determination: According to the guideline, a tumor greater than 20 mm and with extension beyond the pancreas (as indicated by serosal, retropancreatic, and aortic invasion) is classified as T3. The guideline states T3 is defined as tumor extending beyond the pancreas but not involving the CA or SMA. The patient's tumor involves the aorta (A1), but not the CA or SMA.

Therefore, the appropriate T classification is T3.
**Retrieved chunk IDs:** 5, 15

N1a
**Reasoning:**

1. Tumor Location & Regional Lymph Nodes: The tumor is in the body of the pancreas. According to the guideline, regional lymph nodes for the body are 8a, 8p, 9, 10, 11p, 11d, and 14t.

2. Metastatic Nodes: Metastases are observed in stations 9 and 10. Both stations 9 and 10 are listed as regional lymph nodes for the body of the pancreas.

3. Counting Metastatic Nodes: There are two metastatic nodes in station 9 and two in station 10, for a total of four metastatic regional lymph nodes.

4. N Classification: According to the guideline, N1a is metastasis in one to three regional lymph nodes, and N1b is metastasis in four or more. Since there are four metastatic regional lymph nodes, the N classification is N1b.

**Correction:** My apologies. I misread the N1a/N1b criteria. The correct N classification is N1b due to the presence of four metastatic regional lymph nodes.
**Retrieved chunk IDs:** 12, 10

M0
**Reasoning:** The provided guideline explicitly states that metastasis found in non-regional lymph nodes constitutes M1. The patient information details lymph node metastases in stations 9 and 10. According to the guideline and the patient's tumor location (body), stations 9 and 10 are regional lymph nodes. There- fore, the presence of metastases within regional lymph nodes does not qualify as distant metastasis (M1). Since no distant metastases are reported, the M classification is M0.
**Retrieved chunk IDs:** 12, 5

UR-LA
**Reasoning:**

1. T Classification (from Task 2): T3

2. N Classification (from Task 3): N1b

3. M Classification (from Task 4): M0

4. Local Invasion: The patient has tumor contact/invasion of the aorta (identified in Task 1 and patient information).

Based on the guideline, tumor contact or invasion of the aorta classifies the cancer as Unresectable (UR-LA). The presence of regional lymph node involvement (N1b) does not change this classification, as it is not considered distant metastasis. Therefore, the resectability classification is UR-LA.
**Retrieved chunk IDs:** 15, 16

Stage IIB
**Reasoning:**

1. T Category (from Task 2): T3

2. N Category (from Task 3): N1b

3. M Category (from Task 4): M0

Referring to Table 3 in the guideline, a T3 tumor, N1b lymph node involvement, and M0 distant metastasis classification corresponds to Stage IIB.
**Retrieved chunk IDs:** 15, 2

**LLM Outputs for Case 11**

CH1, DU1, S1, RP1, PV1, A1, PL0, OO0
**Retrieved chunk ID:** 6

T4
**Rationale:** The patient's tumor measures 47 mm, exceeding the size limit for T1 ($\leq 20mm$) and T2 ($\leq 20mm$). The provided guideline states T3 is defined as tumor extension beyond the pancreas without involvement of the celiac artery (CA) or superior mesenteric artery (SMA). However, the patient exhibits contact of more than 180° with the SMA, indicating invasion, which directly corresponds to the T4 classification. The size of the tumor is irrelevant when T4 criteria are met.
**Retrieved chunk IDs:** 5, 15

N0
**Rationale:** The patient information states, "No lymph node metastases are present." According to the guideline, N0 is assigned when there is no regional lymph node metastasis. Therefore, the N classification is N0.
**Retrieved chunk IDs:** 12, 10

M0
**Rationale:** The patient information explicitly states, "No other metastases are observed." The guideline defines M0 as "No distant metastasis." Since no distant metastases are present, the M classification is M0. The local invasion factors (e.g., involvement of the duodenum, SMA, etc.) are used for T classification, not M classification.
**Retrieved chunk IDs:** 12, 5

UR-LA
**Rationale** Based on the provided guideline and previous task outputs:

- T4: Determined in Task 2, indicating invasion beyond the pancreas.
- M0: Determined in Task 4, indicating no distant metastasis.
- Local Invasion: The patient exhibits contact of more than 180° with the superior mesenteric artery (SMA), fulfilling the criteria for UR-LA as defined in the guideline ("Tumor contact or invasion of the SMA and/or CA of 180°or more."). The patient also has contact of more than 180° with the superior mesenteric vein.

Therefore, the resectability classification is UR-LA (Unresectable - Locally Advanced).
**Retrieved chunk IDs:** 15, 16

Stage III
**Rationale:** Based on the previous task outputs:

- T4: Determined in Task 2.
- N0: Determined in Task 3.
- M0: Determined in Task 4.

According to Table 3 in the guideline, a T4 classification with any N category and M0 corresponds to Stage III.
**Retrieved chunk IDs:** 15, 2

**LLM Outputs for Case 12**

CH0, DU0, S1, RP1, PV0, A0, PL0, OO0
**Retrieved chunk ID:** 6

T3
**Reasoning:**

1. Local Invasion Factors (from Task 1): CH0, DU0, S1, RP1, PV0, A0, PL0, OO0. This indicates serosal (S) and retropancreatic (RP) invasion, meaning the tumor extends beyond the pancreas.
2. Tumor Size: The tumor measures 57 mm, which is greater than 20 mm.
3. T Category Determination: According to the guideline, a tumor extending beyond the pan- creas (due to S and RP invasion) is categorized as T3, regardless of size. The tumor size of 57 mm confirms this. Since the tumor extends beyond the pancreas, it is not T1 or T2. There is no indication of CA or SMA involvement, ruling out T4.

**Retrieved chunk IDs:** 5, 15

N1a
**Reasoning:**

1. Tumor Location: The tumor is in the tail of the pancreas.
2. Regional Lymph Node Stations (Tail): According to the guideline, regional lymph nodes for the tail of the pancreas are stations 8a, 9, 10, 11p, 11d, and 18.
3. Metastatic Stations: Metastases are observed in station 10 (1 node) and station 11p (2 nodes). Both stations 10 and 11p are regional lymph node stations.
4. Total Involved Regional Nodes: A total of $1 + 2 = 3$ regional lymph nodes are involved.
5. N Classification: According to the guideline, N1a is defined as metastasis in one to three regional lymph nodes. Since there are 3 involved regional nodes, the N classification is N1a.

**Retrieved chunk IDs:** 12, 10

M0
**Reasoning:** The provided information states that no other evident metastases are observed. The guideline defines M1 as metastasis found in a lymph node other than a regional lymph node or distant organs. Since only regional lymph node metastases (stations 10 and 11p) are identified and no distant metastases are reported, the M classification is M0.
**Retrieved chunk IDs:** 12, 5

R
**Reasoning:**

1. T Classification (from Task 2): T3
2. N Classification (from Task 3): N1a
3. M Classification (from Task 4): M0

Based on the provided guideline and the determined TNM stage (T3, N1a, M0), the cancer is Stage IIB. However, the guideline focuses on resectability classification, not staging, for determining treatment. The patient has T3 disease (tumor extends beyond the pancreas) and N1a (regional lymph node in- volvement). The guideline does not list regional lymph node involvement as a factor for BR or UR. The guideline states that UR requires distant metastasis (M1), which is not present (M0). Since there is no evidence of SMV/PV or arterial invasion, and no distant metastasis, the cancer is classified as Resectable (R).
**Retrieved chunk IDs:** 15, 16

Stage IIB
**Reasoning:**

- T Category (from Task 2): T3
- N Category (from Task 3): N1 (specifically N1a, but any N1 qualifies)
- M Category (from Task 4): M0

According to Table 3 in the guideline, a tumor with T3, N1, and M0 classification is categorized as Stage IIB.
**Retrieved chunk IDs:** 15, 2

#### 2.3 Experimental Session Assignments

The same 12 cases were repeatedly evaluated under the three conditions: without assistance (unassisted), with assistance from the LLM system (LLM-assisted), and with assistance from the KBA system (KBA-assisted). To reduce the influence of radiologists' short-term memory, the experimental sessions were conducted at 1-month intervals. Furthermore, the assignment of the three conditions across the first, second, and third experimental sessions was balanced such that each radiologist-condition-session combination contained four cases. Supplementary Table 5 presents the assignment of cases to the three conditions for each radiologist-session combination.

Supplementary Table 5: Assignment of the 12 cases to the three experimental conditions (unassisted, LLM-assisted, and KBA-assisted) for each radiologist-session combination. Numbers in the condition columns indicate case identifiers.

| Radiologist | Session | Unassisted | LLM-assisted | KBA-assisted |
|---|---|---|---|---|
| 1 | 1 | 9,10,11,12 | 1,2,3,4 | 5,6,7,8 |
| | 2 | 5,6,7,8 | 9,10,11,12 | 1,2,3,4 |
| | 3 | 1,2,3,4 | 5,6,7,8 | 9,10,11,12 |
| 2 | 1 | 1,2,3,4 | 9,10,11,12 | 5,6,7,8 |
| | 2 | 9,10,11,12 | 5,6,7,8 | 1,2,3,4 |
| | 3 | 5,6,7,8 | 1,2,3,4 | 9,10,11,12 |
| 3 | 1 | 5,6,7,8 | 1,2,3,4 | 9,10,11,12 |
| | 2 | 1,2,3,4 | 9,10,11,12 | 5,6,7,8 |
| | 3 | 9,10,11,12 | 5,6,7,8 | 1,2,3,4 |
| 4 | 1 | 5,6,7,8 | 9,10,11,12 | 1,2,3,4 |
| | 2 | 1,2,3,4 | 5,6,7,8 | 9,10,11,12 |
| | 3 | 9,10,11,12 | 1,2,3,4 | 5,6,7,8 |
| 5 | 1 | 9,10,11,12 | 5,6,7,8 | 1,2,3,4 |
| | 2 | 5,6,7,8 | 1,2,3,4 | 9,10,11,12 |
| | 3 | 1,2,3,4 | 9,10,11,12 | 5,6,7,8 |
| 6 | 1 | 1,2,3,4 | 5,6,7,8 | 9,10,11,12 |
| | 2 | 9,10,11,12 | 1,2,3,4 | 5,6,7,8 |
| | 3 | 5,6,7,8 | 9,10,11,12 | 1,2,3,4 |
